# Impact of Effective Refractory Period Personalization on Arrhythmia Vulnerability in Patient-Specific Atrial Computer Models

**DOI:** 10.1101/2024.06.06.24308556

**Authors:** Patricia Martínez Díaz, Albert Dasí, Christian Goetz, Laura Unger, Annika Haas, Armin Luik, Blanca Rodríguez, Olaf Dössel, Axel Loewe

## Abstract

**Background and Aims:** The effective refractory period is one of the main electrophysiological properties governing arrhythmia maintenance, yet effective refractory period personalisation is rarely performed when creating patient-specific computer models of the atria to inform clinical decision-making. The aim of this study is to evaluate the impact of incorporating clinical effective refractory period measurements when creating *in silico* personalised models on arrhythmia vulnerability.

**Methods:** Clinical effective refractory period measurements were obtained in seven patients from multiple locations in the atria. The atrial geometries from the electroanatomical mapping system were used to generate personalised anatomical atrial models. To reproduce patient-specific refractory period measurements, the Courtemanche cellular model was gradually reparameterised from control conditions to a setup representing atrial fibrillation-induced remodelling. Four different modelling approaches were compared: homogeneous (A), heterogeneous (B), regional (C), and continuous (D) distribution of effective refractory period. The first two configurations were non-personalised based on literature data, the latter two were personalised based on patient measurements. We evaluated the effect of each modelling approach by quantifying arrhythmia vulnerability and tachycardia cycle length. We performed a sensitivity analysis to assess the influence of effective refractory period measurement uncertainty on arrhythmia vulnerability.

**Results:** The mean vulnerability was 3.4±4.0%, 7.7±3.4%, 9.0±5.1%, 7.0±3.6% for scenarios A to D, respectively. The mean tachycardia cycle length was 167.1±12.6ms, 158.4±27.5ms, 265.2±39.9ms, and 285.9±77.3ms for scenarios A to D, respectively. Incorporating perturbations to the measured effective refractory period in the range of 2, 5, 10 and 20ms, had an impact on the vulnerability of the model of 5.8±2.7%, 6.1±3.5%, 6.9±3.7%, 5.2±3.5%, respectively.

**Conclusion:** Increased dispersion of the effective refractory period had a greater effect on reentry dynamics than on mean vulnerability values. The incorporation of personalised effective refractory period in the form of gradients had a greater impact on vulnerability than had a homogeneously reduced effective refractory period. Effective refractory period measurement uncertainty up to 20ms slightly influences arrhythmia vulnerability. Electrophysiological personalisation of atrial *in silico* models appears essential and warrants confirmation in larger cohorts.

## 1. Introduction

Refractoriness is an electrophysiological property that characterises the response of cardiac tissue to premature stimulation. Shortened cardiac refractoriness promotes sustained re-entrant activity [1] and can be assessed during electrophysiological (EP) studies following the extra stimulus S1S2 pacing technique, where a train of S1 stimuli is given at a certain cycle length followed by a premature S2 stimulus [2]. The effective refractory period (ERP) can then be defined as the longest S1S2 interval that fails to generate a capture in the tissue. Refractory period can only be determined at one region at a time (between stimulus and measurement locations); thus, multiple measurements are necessary for estimations of spatial distribution [1].

Clinical and pre-clinical investigations have demonstrated heterogeneous refractoriness properties across different atrial regions, which also vary from patient to patient [3–5]. During atrial fibrillation (AF), high stimulation frequencies induce electrical remodelling, resulting in shortened action potential duration (APD) and ERP [6]. However, contrary to the belief that prolonged exposure to AF always shortens ERP, patients with persistent AF may exhibit longer ERP due to the presence of atrial dilatation [7]. So, the overall contribution of refractoriness to increased reentrant inducibility remains unclear.

A common theory explaining the existence of AF postulates that both a trigger and a vulnerable substrate are necessary for the initiation and maintenance of AF [8]. Ectopic activity from the sleeves of the pulmonary veins (PV) is the most frequent form of AF triggers [9]. Non-PV triggers have been identified in the crista terminalis (CT), the interatrial septum, the left atrium (LA) posterior wall, the LAA (LAA), the ligament of Marshal, the superior vena cava (SVC), and the coronary sinus; yet their precise role in initiating AF remains uncertain [10]. The vulnerable substrate refers to the presence of atrial fibrosis and dilatation caused by electrical and structural remodelling, and changes in the extracellular matrix (e.g., fibrosis and adipose tissue infiltration, inflammation, atrial dilatation, etc.) [11]. The presence of electrical heterogeneity, such as regional variations in conduction velocity (CV), APD, and ERP, can sustain AF on vulnerable substrates [12]. However, it is still challenging to characterize the vulnerable substrate in a clinical or experimental setting. Understanding the interplay between electrophysiological and structural factors, and how their regional distribution (heterogeneity) influences arrhythmia maintenance remains a complex task in cardiac electrophysiology research.

In this sense, patient-specific atrial computational models provide a robust framework for studying, under controlled conditions, the integrated effect of substrate features unique to each patient and their impact on arrhythmia vulnerability [13–15]. The creation of patient-specific computer models of the atria typically involves anatomical personalization using image data obtained from MR or CT scans or electroanatomic mapping systems (EAMS). Electrophysiological personalization is rarely performed since patient electrophysiological data is usually not available beforehand [16]. Some studies have conducted personalization of atrial electrophysiology, by fitting model parameters to patient clinical data [13, 16–19]. Their findings suggest that personalised electrophysiological parameter values vary among patients and differ from standardised literature parameter values. Nevertheless, the effect of incorporating patient-specific clinical ERP measurements on arrhythmia vulnerability has not yet been assessed. In this work, we investigate the role of incorporating personalised ERP values from various clinical measurements on the *in silico* assessment of arrhythmia vulnerability.

## 2. Methods

### 2.1. Electrophysiological Study

Six patients with a history of AF and prior pulmonary vein isolation (PVI) and one patient with atrial flutter (AFl) underwent an electrophysiological study. Patients gave written informed consent, and the ethics committee of Städtisches Klinikum Karlsruhe gave ethical approval for this work. Electroanatomic maps during sinus rhythm were generated using the Rhythmia 3D mapping system (Boston Scientific, USA). For patients with prior PVI, LA mapping was conducted, whereas for the patient with AFl, the RA (RA) was mapped. ERP measurements were obtained from multiple locations in the atria (5.7±1.4 measurements) following an S1S2 protocol with seven S1 stimuli at a basic cycle length of 500 ms and an S2 stimulus with intervals between 300 and 200 ms, decreasing by 10 ms until loss of capture. ERP measurements were taken in different anatomical regions such as the anterior wall, posterior or lateral wall, appendage and at least one PV in the case of LA geometries. A representative endocardial trace of the stimulation protocol is shown in Figure 1A. ERP was defined as the longest S1S2 interval without capture. To characterise the patient-specific fibrotic substrate, low voltage areas (*<* 0.5 mV) were identified from bipolar voltage maps.

**Figure 1:**
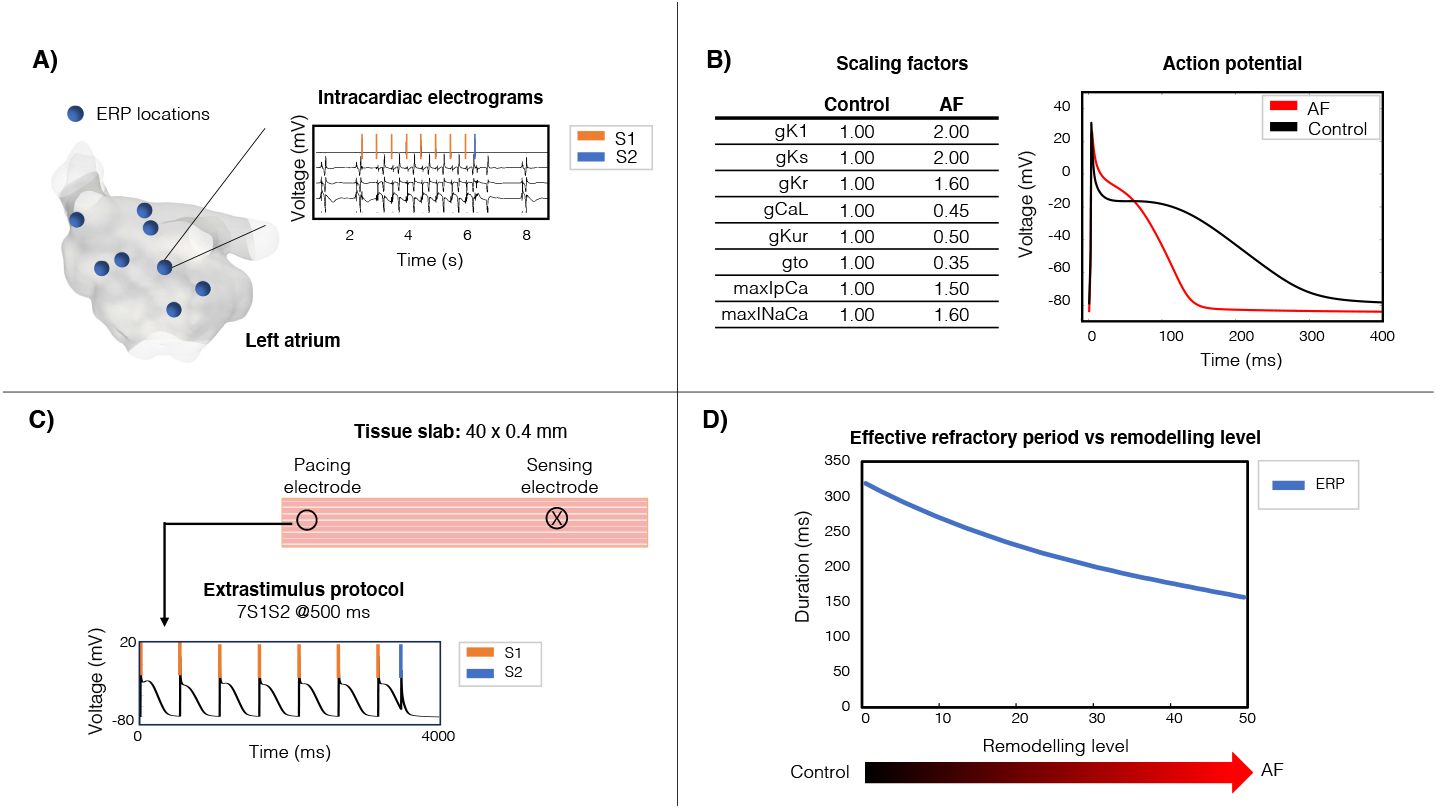
*In silico* representation of clinical patient-specific effective refractory period (ERP). A) The ERP was measured in multiple locations in the atria with an S1S2 protocol. B) Scaling of the maximum ion channel conductances of the Courtemanche cellular model from control to AF. C) Pacing protocol in 1D tissue slab. D) ERP with respect to remodelling level. AF: effective refractory period.

### 2.2. Patient-Specific Anatomical Modelling

The atrial anatomy derived from the EAMS was utilized to generate personalised simulation-ready bilayer meshes. The seven bilayer meshes were created using AugmentA [20] including rule-based anatomical annotations and fibre orientations. For the LA models, the LAA, mitral valve (MV), PV, and left Bachmann’s bundle (BB), were automatically annotated; for the RA model, the tricuspid valve (TV), right atrial appendage (RAA), SVC, inferior vena cava (IVC), pectinate muscles (PM), right BB, and CT. An open-source Python-based algorithm was used to subdivide the meshes into anatomical regions: anterior wall, septal wall, posterior wall, lateral wall, inferior wall, appendage, for the RA and LA accordingly [21].

### 2.3. Atrial Electrophysiology Modelling

Electrical propagation in the atria was modelled using the monodomain equation and simulated with openCARP [22]. Anisotropy in different parts of the atria were modelled as described in [16]. CV was doubled in the CT and tripled in the PM and in the BB [23]. As both ERP and CV influence reentry maintenance [18], our aim was to identify the CV at which vulnerability was highest. We tuned the longitudinal monodomain conductivity to achieve a mean CV of 0.3, 0.5, and 0.7 m/s in the bulk myocardium, for each patient-specific model. A CV of 0.3 m/s, as shown in Figure 2, exhibited the highest number of inducible points and was therefore selected for further vulnerability assessments.

**Figure 2:**
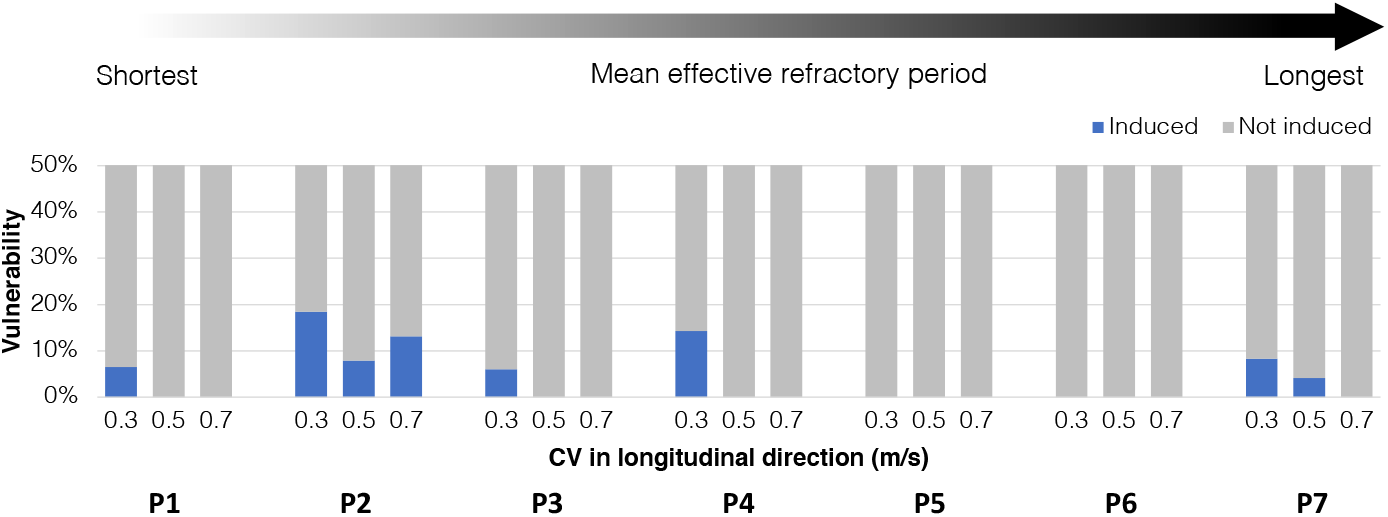
Comparison of arrhythmia vulnerability in scenario B with non-personalized heterogeneous ERP distribution. A CV of 0.3 m/s revealed a higher number of inducing points in the control scenario. CV: conduction velocity, ERP: effective refractory period.

### 2.4. Patient-Specific ERP modelling

To reproduce patient-specific clinical ERP *in silico*, the maximum conductances of key ionic channels affecting AP morphology of the established Courtemanche cellular model [24] were modified from control conditions to a setup leading to changes in the action potential in line with patients with persistent AF [23]. We modified the maximum conductances of the inward rectifier K^+^ current (GK1), the ultrarapid (GKur), rapid (GKr), and slow delayed-rectifier K^+^ currents (Gs), the L-type Ca^2+^ current (GCaL), the transient outward K^+^ current (Gto), the sarcoplasmic Ca^2+^ pump current (IpCa), and the Ca^2+^/Na^+^ exchanger (maxINaCa). The scaling factors can be found in Table1B. The ion channel conductances were linearly scaled to generate a set of 50 different cellular models with gradually increasing remodelling levels. To obtain the ERP of each cellular model, we generated a 1D tissue slab with a size of 40 × 0.4 mm and performed a virtual S1S2 pacing protocol (Figure 1 C-D).

### 2.5. Generation of ERP Scenarios

To assess the role of ERP personalization, we generated four scenarios: homogeneous (A), heterogeneous (B), regional (C), and continuous (D) ERP distribution (Figure 3. The first two configurations were non-personalised based on literature data, the latter two were personalised based on patient measurements. In scenario A, the same cellular model corresponding to AF-induced remodelling [23] was applied to the whole atrium. In scenario B, anatomical structures had individual cellular model variants with specific ERP assigned based on literature data [16]. For scenario C, all nodes in anatomical regions were assigned distinct cellular models with the ERP value matching the spatially closest available clinical measurement. In case of multiple measurements present in the same region, the average ERP value was considered for the whole region. In scenario D, the measured ERPs were assigned to the corresponding catheter tip positions and then continuously mapped to the whole surface by Laplacian interpolation [25]. The measuring points were defined as boundary conditions, so that the ERP values never exceeded the measured ERP range. The ion channel conductances for each individual mesh node were then adjusted accordingly to match the interpolated ERP. The four personalization scenarios are shown in Figure 1.

**Figure 3:**
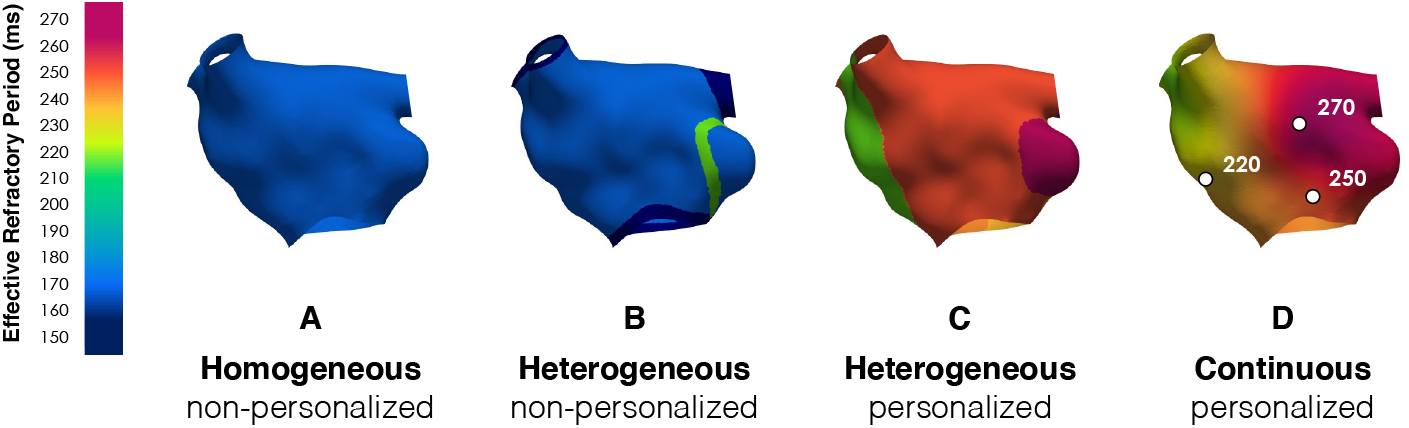
Scenarios for the evaluation of ERP personalization. Homogeneous (A) with non-personalised ERP based on literature data [23], heterogeneous (B) with different non-personalised ERP based on anatomical structures [16], regional (C) with personalised ERP divided into anatomical regions, and continuous (D) with personalised ERP from interpolated measurements. White points denote catheter tip locations where the pacing stimulus was delivered.

### 2.6. Patient-Specific Substrate Modelling

Substrate was incorporated into the meshes based on the identification of low voltage area (LVA). To distinguish between scar tissue from PVI and native fibrosis, we defined scar regions with voltage¡0.1 mV and fibrosis regions between 0.1 and 0.5 mV [12]. To assess the influence of scar and fibrosis on arrhythmia inducibility, we created four additional scenarios, namely A2, D2, A3 and D3. The first two scenarios, A2 and D2, included scar tissue and native fibrosis.

Scenario A2 had the same ERP as scenario A (homogenous) and scenario D2 corresponding to same ERP personalization as scenario D (continuous). Lastly, to model a stage before PVI, we generated scenarios A3 and D3 including only native fibrosis regions, and scar tissue was modelled as healthy. For regions defined as native fibrosis, 30% of the elements were set to non-conductive with *σ* = 10^−7^ S/m to represent electrical myocyte decoupling, while the remaining 70% were set to have electrical remodelling in response to cellular inflammation. For scar regions, all elements were set to be non-conductive [26, 27].

### 2.7. Vulnerability Assessment

Arrhythmia vulnerability was assessed by virtual S1S2 pacing at different locations in the atria separated by an average distance of 2 cm [28]. The vulnerability ratio was defined as the number of inducing points divided by the number of stimulation points. Stimulation points locations remained consistent among scenarios. Transmembrane voltage traces were recorded for 1 s for each reentry at the inducing stimulus location. We determined the tachycardia cycle length (TCL) of the reentries by calculating the average between peaks of d*V* /d*t*.

### 2.8. Sensitivity Analysis

To study the influence of uncertainty in ERP measurements, we conducted a sensitivity analysis by including perturbation in ERP measurements in the ranges of ±2, ±5, ±10 and ±20 ms, randomly drawn from a uniform distribution. We generated 10 perturbation sets for each perturbation range, resulting in a total of 40 new perturbed ERP sets; a separate random value was drawn for each measured ERP. Finally, we generated a new interpolated map using the perturbed ERP sets. Due to the high computational cost of the vulnerability assessment (15±2.4 min per stimulation point, utilizing 4 nodes × 40 CPU cores with Intel Xeon Gold 6230 2.1Ghz), the sensitivity analysis was limited the assessment to patient P3 model, which showed the highest vulnerability in the LA model cohort.

### 2.9. Statistical Analysis

The data are presented as mean±SD. We used a two-sample t-test to determine statistical significance between the sample means. P-values ¡0.05 were considered significant.

### 2.10. Data Availability

Bilayer models and source code to reproduce the simulated reentries are accessible under open licenses [29].

## 3. Results

Patient characteristics are outlined in Table 1. The overall mean clinically measured ERP was 254.0 ± 32.7 ms. The dispersion of the ERP measurements is shown in the boxplot in Figure 4A. The ERP distribution maps for each patient is illustrated in Figure 4B. Bipolar voltage maps for each patient are shown in Figure 5A. LVA accounted for 42.8 ± 16.4 % of the atrial surface. The amount of fibrosis, scar, and healthy tissue for each patient are shown in Figure 5B.

**Table 1:**
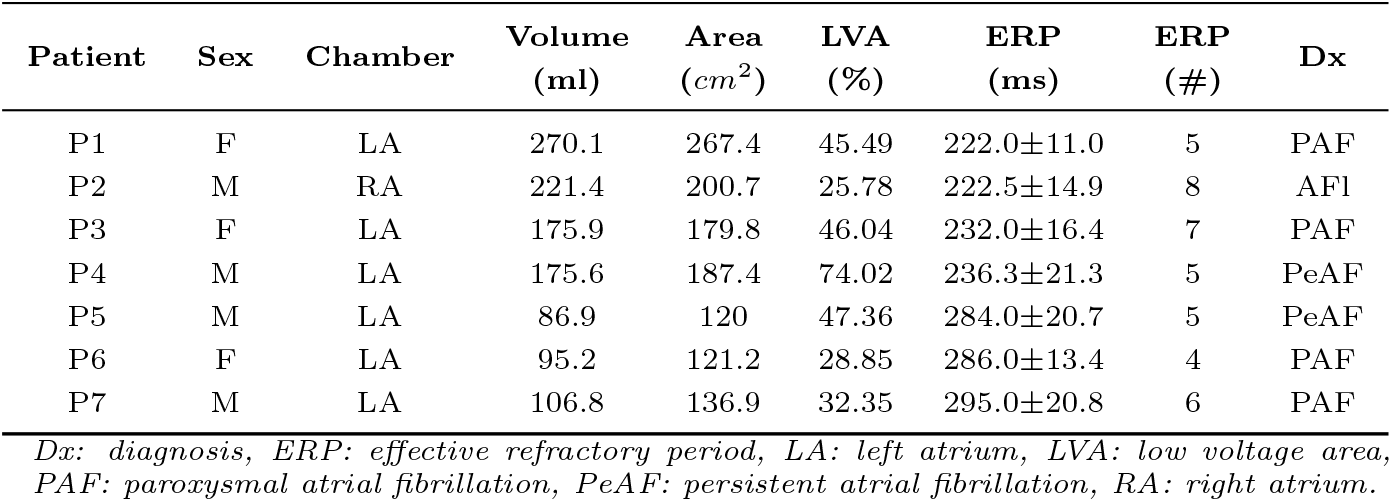
Clinical characteristics of patient cohort.

**Figure 4:**
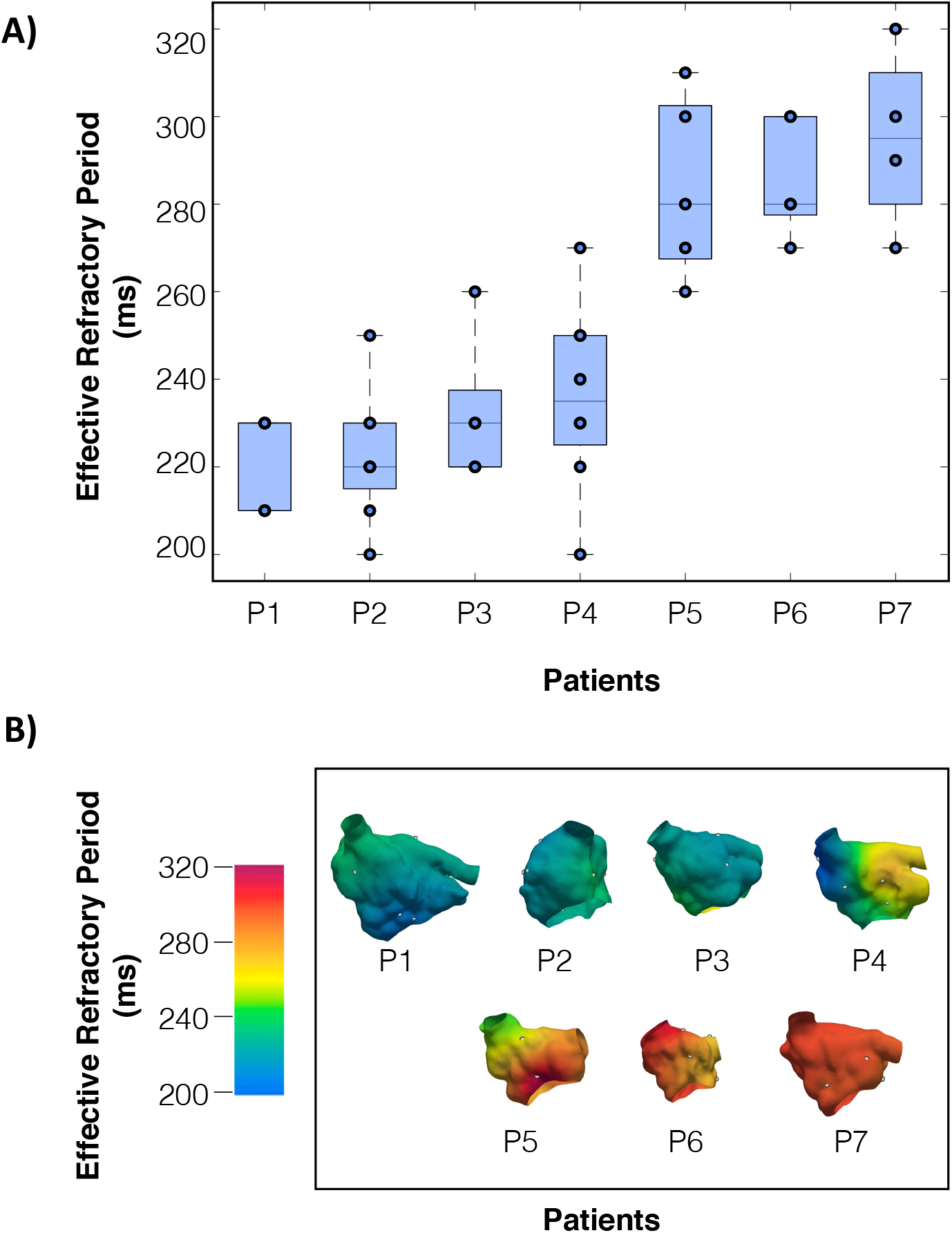
Distribution of effective refractory period (ERP). 1) Boxplots show the dispersion of ERP measurements for each patient, where the points represent each individual measurement. 2) ERP distribution map generated from interpolated clinical measurements from an anterior view.

**Figure 5:**
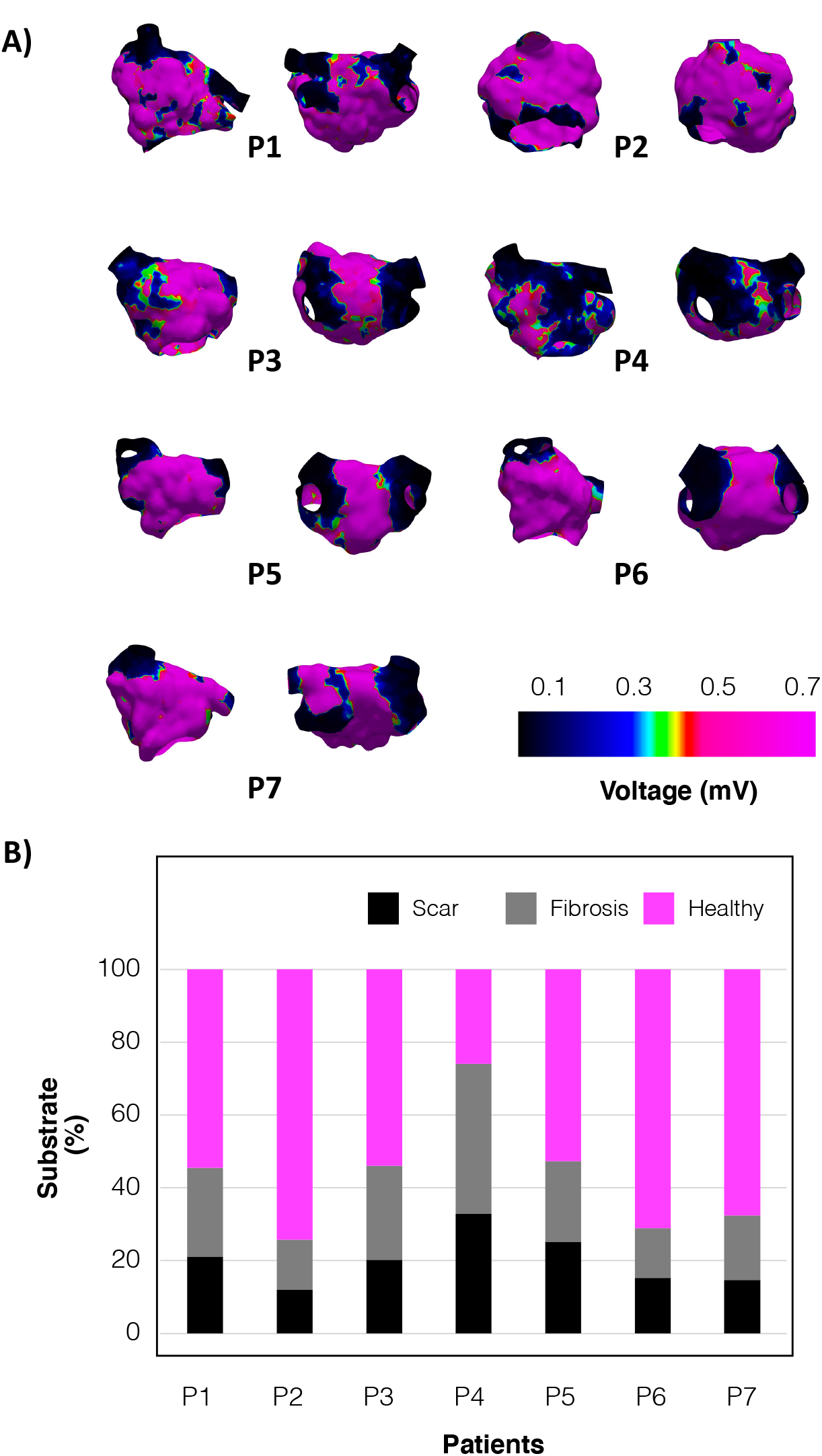
Substrate distribution based on the identification of low voltage areas. A) Bipolar voltage maps from an anterior view. B) Percentage of substrate defined as scar (≤ 0.1 mV) shown in black, fibrotic regions (0.1-0.5 mV) in grey and healthy tissue (≥ 0.5 mV) in magenta.

*In silico* ERP of tissue patch models varied from 320 ms in the control state to 157 ms in the AF remodelling state. Non-personalized scenarios had a shorter ERP and reduced dispersion with ERP in the range of 158.9 ± 5.3 ms for non-personalized scenarios, and 254.0 ± 32.7 ms for personalized scenarios. From a total of 214 stimulation points (30.6 ± 8.9 stimulation points per patient), 61 simulated reentries were induced across the four scenarios without fibrotic substrate, with individual counts of 7, 18, 20, and 16 reentries for scenarios A, B, C, and D, respectively. Vulnerability values are shown in Figure 6. The vulnerability for scenario A was 3.4 ± 4.0%, 7.7 ± 3.4% for scenario B, 9.0 ± 5.1% for scenario C and 7.0 ± 3.6% for scenario D. The mean TCL was 167.07 ± 12.58 ms for scenario A, 158.42 ± 27.52 ms for scenario B, 265.17 ± 39.87 ms for scenario C and 285.88 ± 77.31 ms for scenario D, as shown in Figure 7.

**Figure 6:**
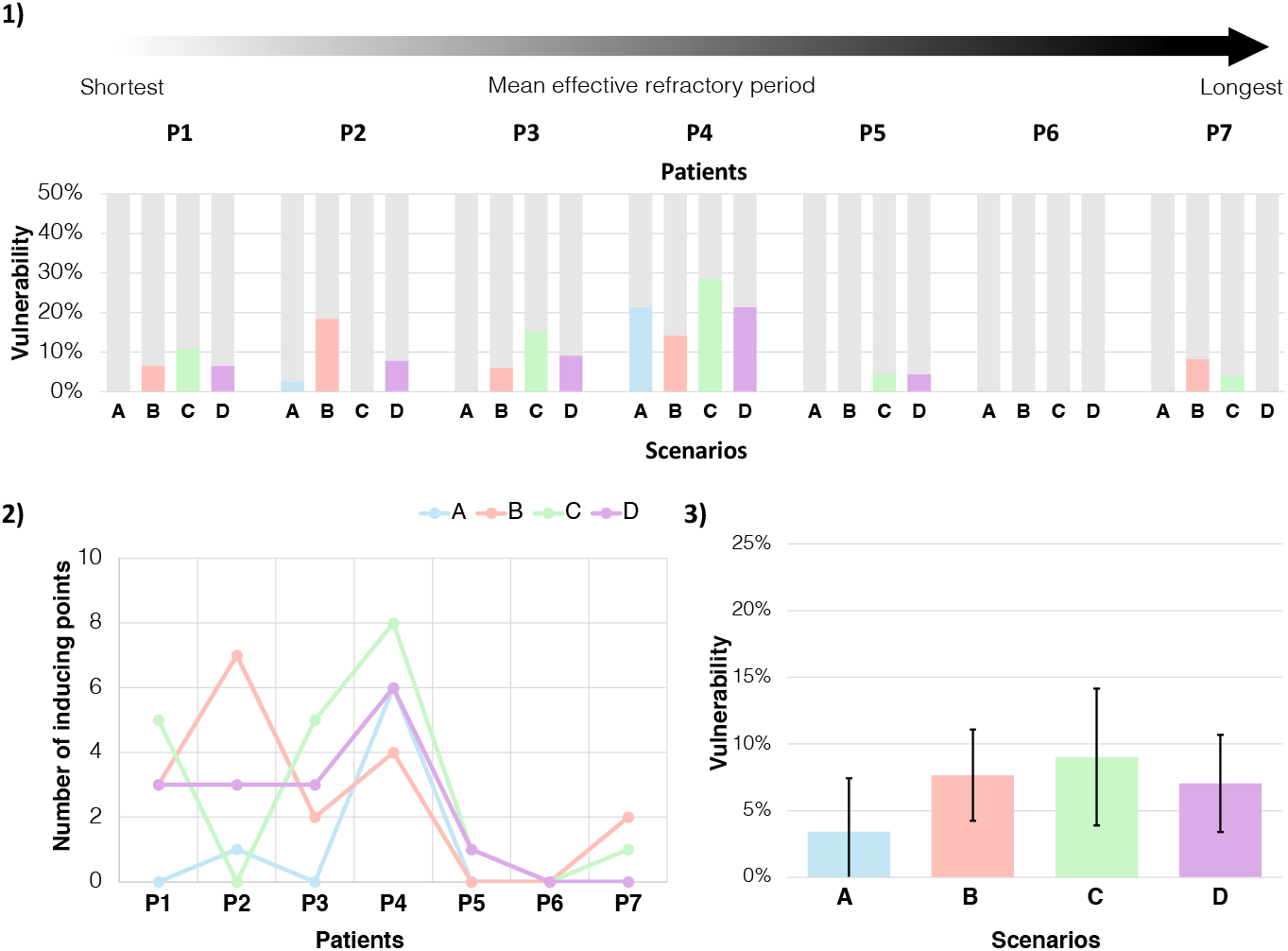
Comparison of arrhythmia vulnerability and tachycardia cycle length among four scenarios. 1) Vulnerability for each patient in four personalization scenarios without fibrotic substrate 2) Number of inducing points for each patient. 3) Mean vulnerability, bars indicate standard deviation. P1-P7 indicate individual patients. Scenarios are defined as A: homogeneous, B: heterogeneous, C: regional, and D: continuous ERP distribution.

**Figure 7:**
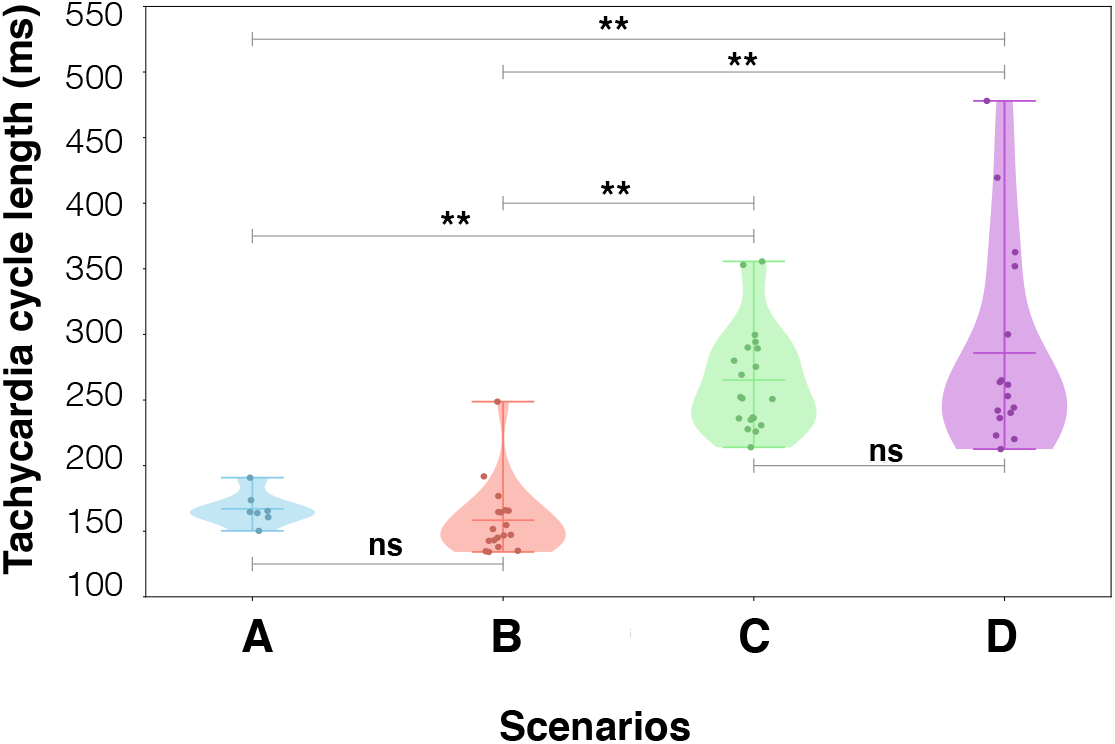
Tachycardia cycle length of induced reentries. Each individual point represents one reentry Conduction velocity in the bulk myocardium was set to 0.3 m/s in longitudinal direction. **: p*<*0.05, ns: not statistically significant. Scenarios are defined as A: homogeneous, B: heterogeneous, C: regional, and D: continuous ERP distribution.

We assessed the impact of incorporating fibrotic substrate informed by LVA into the model along with ERP personalization on arrhythmia vulnerability. Given that most patients had undergone previous PVI, we compared vulnerability with and without ablation scars defined by regions where bipolar amplitude was *<* 0.1 mV. To avoid additional confounding factors, we compared scenario A (homogeneous non-personalized) and scenario D (continuous with personalized ERP) without fibrosis with their respective counterparts with fibrosis and scar A2 and D2 (Figure 8). Incorporating fibrosis and scar resulted in a vulnerability of 11.3 ± 7.28% and 3.93 ± 3.33% for A2 and D2, respectively. Incorporating only fibrosis without scar resulted in a vulnerability of 47.54 ± 31.96% and 39.4 ± 30.31% for A3 and D3, respectively. Area reduction due to PVI decreased vulnerability in A2 and D2 vs A3 and D3 (Figure 8). The magnitude of the difference between A3 and D3 varied among patients, with some experiencing small differences in vulnerability, e.g., P2 and P4, while others showing a bigger difference, e.g., P1 and P7. On average, lower mean ERP in A3 in the presence of native fibrosis without scar resulted in higher vulnerability compared to D3.

**Figure 8:**
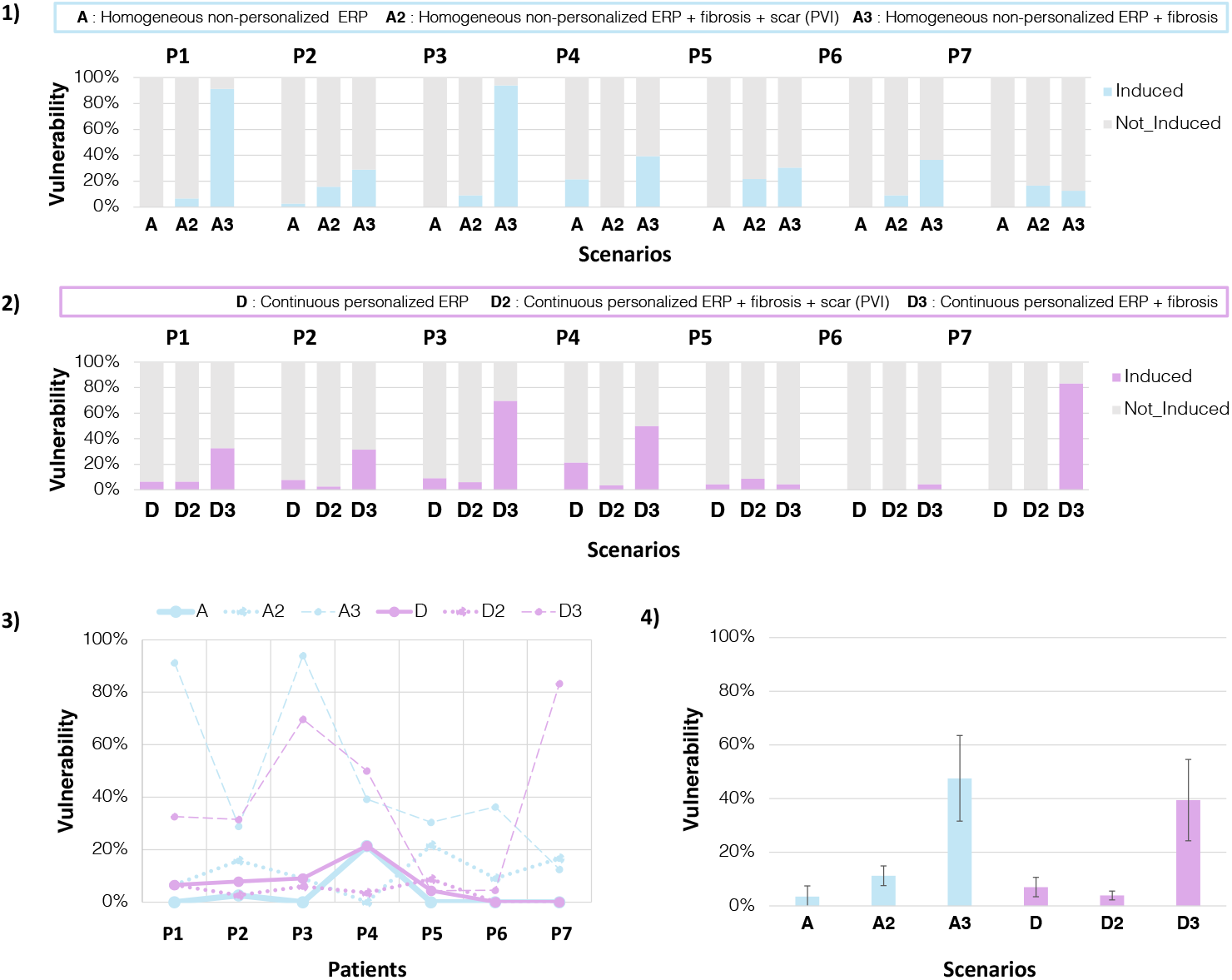
Comparison of fibrotic substrate modelling and ERP personalization between scenario A (homogeneous non-personalised ERP) and scenario D (continuous with personalised ERP). 1) Vulnerability for scenario A and two LVA substrate modelling scenarios: A2 with scar (*<*0.1 mV) and fibrosis (*<*0.5 mV), and A3 as a state before PVI with only fibrosis (*>*0.1 mV). 2) Vulnerability for scenario D and two LVA substrate modelling scenarios: D2 with personalised continuous ERP + with scar (*<*0.1 mV) and fibrosis (*<*0.5 mV) and D3 as a state before PVI with personalised continuous ERP and only fibrosis (*>*0.1 mV). 3) Comparison of six scenarios for each patient. 4) Mean vulnerability per scenario. Bars denote standard deviation. ERP: effective refractory period, LVA: low voltage area. P1-P7 indicate individual patients.

Incorporating perturbations to the measured ERP in the sensitivity analysis slightly impacted the vulnerability of the model from 9.1% to 5.8 ± 2.7%, 6.1 ± 3.5%, 6.9 ± 3.7%, and 5.2 ± 3.5%, observed for perturbations in the range of ±2, ±5, ±10 and ±20 ms, respectively (Figure 9).

**Figure 9:**
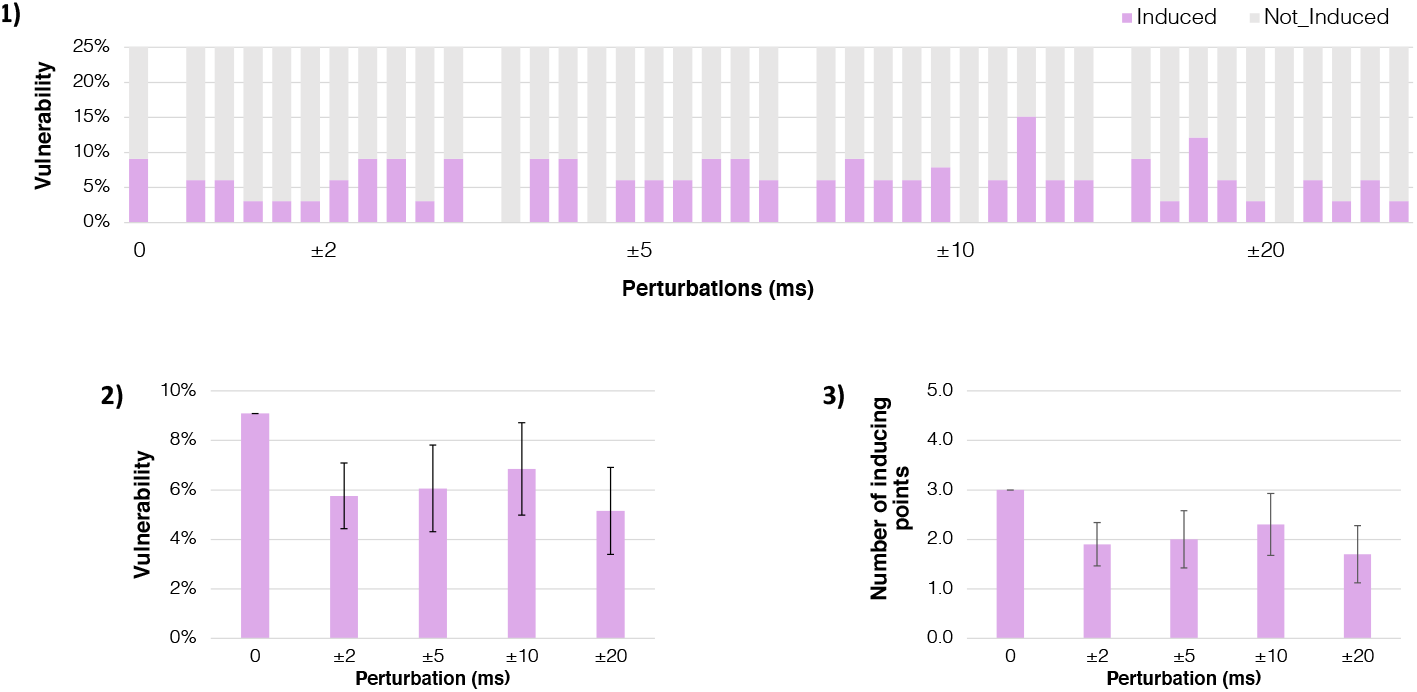
Sensitivity analysis for arrhythmia vulnerability of patient P3 comparing scenario D (personalised with continuous ERP) and perturbed ERP measurements. 1) Vulnerability for each perturbation range. 2) Mean vulnerability. 3) Mean number of inducing points for each perturbation range. Bars indicate standard deviation. Perturbations of ±2, ±5, ±10, and ±20 ms, were randomly drawn, 10 times for each perturbation range, from a uniform distribution. The perturbations were incorporated to each measured ERP value and a new interpolated continuous maps were generated from the perturbed ERP measurements.

## 4. Discussion

In this study, we assessed arrhythmia vulnerability in a cohort of seven patient-specific atrial models, each with information on the distribution of ERP and low voltage substrate. We compared vulnerability across four ERP personalization scenarios: non-personalized homogeneous (A), non-personalized heterogeneous (B), personalized regional (C), and personalized continuous (D) distribution of ERP without substrate. Secondly, we investigated the impact on vulnerability of the interaction between native fibrosis and scar with ERP. Thirdly, we conducted a sensitivity analysis to evaluate the effects of uncertainty in ERP measurements on arrhythmia vulnerability. The four main highlights of our study are: 1) differences in arrhythmia vulnerability between personalised and non-personalised scenarios should be acknowledged, particularly for patients with low ERP, 2) an increased dispersion of the ERP in personalised scenarios had a greater effect on reentry dynamics than on mean vulnerability values, 3) the incorporation of personalised ERP had a greater impact on inducibility than had a homogeneously reduced ERP, with this effect reversing once fibrosis was included, and 4) ERP measurement uncertainty up to 20ms slightly influences arrhythmia vulnerability.

### 4.1. Effect of ERP Personalization on Arrhythmia Vulnerability and Dynamics

Personalised and non-personalised scenarios were different in the mean and dispersion of the ERP. Among all scenarios, the homogeneous non-personalised scenario A had the lowest vulnerability, while the regional personalised scenario C had the highest vulnerability. Heterogeneities in the form of regions in scenario C promote unidirectional blocks, thereby increasing vulnerability, while the homogeneous scenario A makes it less likely to induce reentry even with a shorter ERP [30]. Differences in vulnerability between personalized and non-personalized scenarios were greater in patients with lower ERP (*<* 240 ms), corresponding to P1-P4, with a total of 56 inducing points. In contrast, the remaining three patients (P5-P7) were almost non-inducible, with only five inducing points in total. Decreased inducibility in P5-P7 could be attributed to the reduced atrial conducting size [31]. Thus, differences in vulnerability between personalised and non-personalised scenarios cannot be neglected, particularly for patients with low ERP.

The effect of ERP personalisation becomes more evident when analysing reentry dynamics. There were no significant differences in the TCL of the non-personalised scenarios A and B (167.1±12.6 ms vs. 158.4±27.5 ms, *p* = 0.43), nor were there significant differences between the personalised scenarios C and D (265.2±39.9 ms vs. 285.9±77.3 ms, *p* = 0.31). However, personalised scenarios had significantly slower TCL compared to non-personalised scenarios (*p <* 0.001). This finding suggests that the increased dispersion of the ERP in the personalised scenarios has a greater effect on reentry dynamics than on the absolute value of vulnerability.

### 4.2. Increased ERP Dispersion is Associated with Higher Arrhythmia Vulnerability

Previous clinical and simulation studies have analyzed the effect of ERP and APD dispersion on arrhythmia vulnerability in patients with persistent and paroxysmal AF [7, 32–34]. Dispersion can be defined both spatially and temporally. Spatial dispersion refers to the difference between the maximum and minimum values of ERP measurements [34], while temporal dispersion refers to variation exceeding 5% from the baseline value [33]. In the study of Narayan et al., pacing-induced AF from either the PVs or high RA was always preceded by an increased temporal APD dispersion [33]. In a cohort of 47 patients with paroxysmal AF, ERP was measured in 5 sites in both atria and a higher ERP dispersion was found to be the only clinical predictor of AF inducibility [34]. An interesting finding of this study was that in patients with induced AF, ERP dispersion was similar in those with self-sustained and self-terminated AF. In another cohort of 22 patient-specific biatrial models without personalized ERP, where the substrate was modeled based on late gadolinium enhancement magnetic resonance imaging by applying changes in anisotropy, conduction, and remodeled electrophysiology, the 13 models in which AF was induced had significantly larger APD gradients [35]. In our results, simulations with higher ERP dispersion and lower ERP mean had higher inducibility.

Previous studies have demonstrated that introducing a ±10% homogeneous variation to the baseline APD increases uncertainty in both the quantity and preferred locations of reentrant drivers [36, 37]. Consequently, it is expected that higher variations would also impact reentry inducibility. In our results, the clinical ERP ranged from 222 ms to 295 ms, indicating a variation of 32.9%. When compared to literature-based ERP values (157 ms), the variation between the literature-based value and the maximum observed clinical ERP reaches 87.9%. We conclude that increased inducibility depends on both reduced mean ERP and increased dispersion.

### 4.3. Interaction Between ERP and Substrate Heterogeneities

In current clinical practice, it remains challenging to identify patients for whom PVI will be sufficient to prevent AF recurrence without additional extra-PVI ablation. Six out of seven patients in our cohort had undergone prior PVI, indicating that PVI was ineffective in preventing AF recurrence. It is likely that substrate progression and gaps in PVI promoted AF recurrence [38, 39]. of by the Regional heterogeneities in ERP dispersion are believed to be capable of sustaining AF on vulnerable substrates. Several atrial *in silico* studies have shown that fibrosis regions can anchor or block reentrant drivers [35, 40, 41]. In general, our results showed that fibrosis and scar had a higher impact on vulnerability than ERP. However, the combination of fibrotic substrate and ERP had different effects on vulnerability. Scenarios in which both native fibrosis and scar were considered had lower vulnerability compared to those having native fibrosis only. A possible explanation is the reduced effective atrial size due to PVI scar [31]. We tried to simulate a state prior to PVI in scenarios A3 and D§, although it is likely that native fibrosis based on the identification of regions with voltage *>*0.1 and *<*0.5,**m***V* might not accurately represent the pre-ablation state. On average, the lower mean ERP in A3 in the presence of native fibrosis without scar resulted in higher vulnerability compared to a dispersed ERP distribution as in D3. As substrate areas have a significant impact on model inducibility, further studies should focus on providing a more detailed description of their spatial distribution for informing patient-specific models.

### 4.4. Incorporating Uncertainty to ERP measurements

Measured ERP values depend on the time resolution of the S2 coupling interval, with higher resolution leading to more accurate values. To determine if adding ERP perturbations *<*10 ms would affect vulnerability, we conducted a sensitivity analysis by running 40 additional vulnerability assessments. Our results suggest that variations in the range of 2-20 ms did not markedly change the number of inducing points, and vulnerability remained similar. Indicating that further reductions in the S2 coupling interval ERP would not impact model inducibility. However, these differences in vulnerability might become more pronounced when functional substrate is incorporated. As mentioned before, reentry dynamics are affected when ERP is personalised, rather than inducibility; therefore, future studies should assess the impact of ERP uncertainty on reentry dynamics.

## 5. Limitations

The small sample size can limit the generalisation of our findings. In the optimisation process to adapt the cellular electrophysiology model to measured ERP, we reduced the dimensionality of the parameter set space by constraining the range of variation for each parameter in a linear fashion from normal healthy to changes due to persistent AF. We did not personalise CV distribution. The rate-dependent nature of ERP was not evaluated in our study as clinically measurements of the ERP were only obtained at 500 ms S1 cycle length.

## 6. Conclusions

Incorporation of patient-specific ERP values affects the assessment of AF vulnerability. Differences in arrhythmia vulnerability between personalised and non-personalised scenarios should be acknowledged, particularly for patients with low ERP.An increased dispersion of the ERP in personalised scenarios had a greater effect on reentry dynamics than on mean vulnerability values. The incorporation of personalised ERP had a greater impact on inducibility than had a homogeneously reduced ERP, with this effect reversing once fibrosis was included. ERP measurement uncertainty up to 20 ms slightly influences arrhythmia vulnerability. Functional personalisation of atrial *in silico* models appears essential and warrants confirmation in larger cohorts.

## 7. Funding

This project has received funding from the European Union’s Horizon 2020 research and innovation programme under the Marie Sk-lodowska-Curie grant agreement No 860974. This work was supported by the Leibniz ScienceCampus “Digital Transformation of Research” with funds from the programme “Strategic Networking in the Leibniz Association”. The authors acknowledge support by the state of Baden-Württemberg through bwHPC.

## Notes

### Competing Interest Statement

The authors have declared no competing interest.

### Funding Statement

This project has received funding from the European Unions Horizon 2020 research and innovation programme under the Marie Skłodowska-Curie grant agreement No 860974. This work was supported by the Leibniz ScienceCampus "Digital Transformation of Research" with funds from the programme "Strategic Networking in the Leibniz Association". The authors acknowledge support by the state of Baden-Wuerttemberg through bwHPC.

### Author Declarations

The ethics committee of Staedtisches Klinikum Karlsruhe gave ethical approval for this work.

